# Unifying Vision and Language: Benchmarking End-to-End Transformer Model Against the ClipCap Framework

**DOI:** 10.64898/2026.09.24.26363914

**Authors:** Shreya Anand, M S Dinesh, Andre Dekker, Leonard Wee

**Affiliations:** Department of Radiation Oncology (Maastro), GROW Research Institute for Oncology and Reproduction, Maastricht University Medical Centre+, Netherlands; Clinical Informatics, Advanced Development, Philips Innovation Campus, Embassy Tech Hub, Yelahanka, Bangalore-560064, India

**Keywords:** Vision-language, transformer, Clip, MR cardiac, view labelling

## Abstract

**Background and purpose:** Accurate identification of cardiac MRI volume orientations is essential for reliable image interpretation and for enabling downstream automated analysis pipelines. However, orientation labels and commonly assigned manually, making the process time-consuming and prone to variability. Recent advances in vision-language models offers new opportunities for automated label generation by jointly modeling visual and semantic information.

**Methods:** In this study, we systematically compare a conventional end-to-end transformer-based captioning model with a CLIPCap architecture for automated cardiac MRI view labelling. Both models were evaluated on a dataset of 1184 de-identified cardiac MR images spanning across six clinically relevant orientations: short axis, two-, three-, four-chamber views, left and right ventricular outflow tract views. The caption transformer was trained from scratch, whereas CLIPCap leveraged pretrained CLIP image embeddings combined with lightweight mapping network and a frozen language model.

**Results:** Experimental results demonstrate a substantial performance advantage from CLIPCap, which achieved an overall accuracy of 98% compared to 68% for the caption transformer. CLIPCap consistently delivered high precision and recall across all orientation classes, including those of high clinical importance, while the end-to-end transformer exhibited unstable performance and complete failure in certain views.

**Conclusion:** These findings highlight the benefits of leveraging large-scale multimodal pretrained model for medical image labeling tasks, particularly in data-limited settings. The study suggests that CLIPCap provides a more robust and clinically reliable solution for automated cardiac MRI orientation labeling, supporting its integration into efficient and scalable clinical workflows.

## 1. Introduction

In recent years, the combination of computer vision and natural language processing has witnessed remarkable progress, driven by advances in deep learning techniques and the availability of multimodal datasets. Among the most transformative developments in this space is the emergence of models capable of generating descriptive natural language descriptions from visual inputs, a task known as image captioning. Automatic label generation, which is a subtask of image captioning, has become a critical component of applications related to assistive technologies, especially in the medical domain. The task requires models to not only recognize visual features but also to generate linguistically coherent and semantically relevant labels.

Image captioning has traditionally relied on encoder-decoder architectures using convolutional neural networks (CNNs) and recurrent neural networks (RNNs) [1][2]. However, these models are known to have trouble with long-range dependencies and semantic coherence [3]. Traditional sequence-models using recurrent architectures have now been largely replaced by deep learning approaches that integrate convolutional or transformer-based encoders with powerful language decoders, and this approach has shown to be more promising [3][4].

The introduction of transformer architecture, originally designed for natural language processing, brought a paradigm shift to this domain. Transformers utilize self-attention mechanism to model global dependencies in data, enabling more accurate and contextually rich caption generation. Recent surveys have highlighted the superiority of transformer-based models over traditional approaches, especially in multimodal settings [5]. These models have demonstrated improved performance on benchmark datasets such as MS COCO and Conceptual Captions, using metrics like BLEU, METEOR, CIDEr, and ROUGE [6]. Their ability to integrate visual and linguistic features through attention mechanisms has made them the backbone of state-of-the-art captioning systems.

While transformer-based architectures have significantly advanced the capabilities of image captioning by modeling complex dependencies and enabling richer multimodal interactions, they still rely heavily on supervised learning with curated datasets. This dependency often limits their generalizability across diverse domains and tasks. These constraints of supervised captioning models, led to the development of CLIP (Contrastive Language–Image Pretraining), a model that redefined multimodal learning by aligning visual and textual representations through contrastive learning. CLIP was trained on 400 million image-text pairs collected from the internet using a contrastive learning objective [8]. It employs dual encoders for images and text, aligning them in a shared embedding space. This design allows CLIP to perform zero-shot classification and retrieval tasks without task-specific fine-tuning, making it highly generalizable across domains.

Building on CLIP’s semantic richness, Mokady et al. [9] introduced CLIP-Cap, a lightweight and efficient image captioning model. CLIP-Cap uses CLIP’s image embeddings as a prefix to guide a transformer-based language model (GPT-2) in generating captions. A simple mapping network translates CLIP features into a format compatible with the language model, enabling fluent and context-aware caption generation. CLIPCap bridges CLIP’s vision-language embeddings with a captioning transformer.

Notably, CLIP-Cap achieves competitive results in Conceptual Caption generation without requiring extensive fine-tuning or additional annotations [9]. Its architecture allows both CLIP and GPT-2 to remain frozen during training, significantly reducing computational overhead. This approach exemplifies the shift toward leveraging pretrained multimodal representations for efficient downstream tasks, like label generation. Since CLIP has been pre-trained on natural images to match the image and text in the same embedding space, the amount of data required to fine-tune the model is comparatively less than that required for a Caption Transformer.

The success of transformers and CLIP-based models in general-purpose captioning has catalyzed their adoption in the medical domain. While both architectures have demonstrated success, there has been limited work directly comparing their performance under controlled experimental settings. This paper addresses that gap by conducting a systematic evaluation of a vanilla Transformer-based label generation model and ClipCap model on a medical dataset consisting of MR Cardiac images. These models are trained to generate the orientation details of the MR Cardiac volume. We compare them not only in terms of accuracy but also with respect to efficiency, robustness, and qualitative label quality, thereby offering insights into their relative strengths and practical trade-offs.

Cardiac Imaging using MRI (Magnetic Resonance Imaging), has proven to be a powerful diagnostic and prognostic tool in cardiovascular domain, as it produces high resolution images without ionizing radiation. Its usefulness spans across analyzing structural, functional, perfusion and tissue characterization qualities of the heart. Having orientation details of the acquired volume, which provide the spatial positioning of the image and the heart’s anatomical structure in a 3D space, promotes accurate visualization, diagnosis and comparison across time points. The orientation is usually entered by a Subject Matter Expert (SME) after acquisition, which is used for further analyses. Analyzing each and every MR cardiac volume manually, and providing the orientation details, is time consuming and also prone to human error. Our proposed method aims to automatically label the orientation of the MR cardiac volume using DL (Deep Learning) based methods.

Having the orientation already present will aid in faster diagnosis and save time and effort for a radiologist. It also offers several additional advantages. Orientation labels allow radiologists and cardiologists to accurately identify which part of the heart is being imaged, enabling the identification of structures such as the septum, lateral wall, apex, and base. They improve the accuracy of image registration and 3D reconstruction, help align follow-up scans for easier comparison, facilitate the use of AI and post-processing tools for correct structure identification and motion tracking, and support image segmentation. Additionally, orientation information enhances interoperability between different medical imaging systems.

Cardiac MRI plays a major in cardiovascular assessment due to its high-resolution depiction of cardiac anatomy and function without ionizing radiation. Orientation labels, typically provided expert readers, are essential for anatomical interpretation, which further enable downstream AI tasks such as identifying structures and quantify cardiac motion. However, manual annotation is time-consuming and susceptible to variation, highlighting the need for automated labeling methods. This setting provides a controlled context in which the comparison of transformer-based caption model with ClipCap is performed. In this work, we systematically evaluate a vanilla transformer model and a CLIP-Cap model for generating orientation labels for MR cardiac volumes. We hypothesize that the CLIP-Cap architecture—owing to its large-scale contrastive pretraining, lightweight mapping networks and reduced training complexity, will demonstrate superior robustness and efficiency, while achieving improved accuracy.

## 2. Methodology

### 2.1 Dataset

The dataset used for this experiment consists of deidentified, non-contrast MR Cardiac volumes with 6 orientations with institutional approval. They are T1-weighted images acquired from 1.5T and 3T, multi-vendor scanners. The orientations include Short Axis (SA), 2 chambers (2CH), 3 chambers (3CH) and 4 chambers (4CH), Left Ventricular Outflow Tract (LVOT), Right Ventricular Outflow Tract (RVOT) as in Figure 1. The mid slices in the volume are analyzed, in which the orientation is clearly visible and the appropriate orientation labels are provided for the volume. A total of 1184 images were used in the experiment, with an 80:10:10 split for the training, validation, and test datasets. These were stratified across the respective classes, with 2CH containing 201 images, 3CH 136 images, 4CH 355 images, LVOT 176 images, RVOT 97 images and SA 219 images.

**Figure 1:**
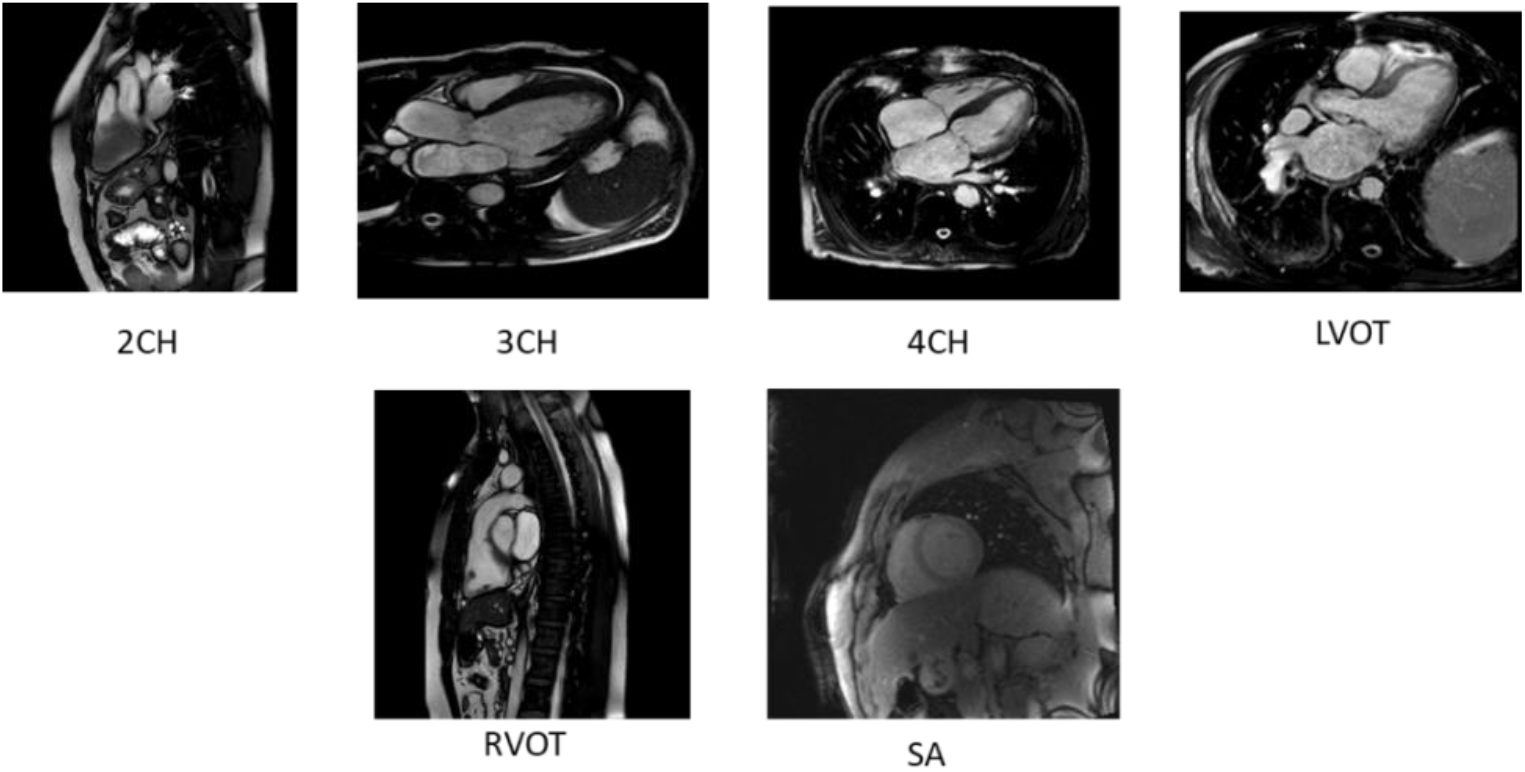
Different orientational views in the MR Cardiac dataset

#### Each of the orientational views has a specific intention in terms of diagnosing pathologies

- 2CH: Captures the left atrium and left ventricle along with mitral valve motion. Provides insights into left ventricular function.
- 3CH: Shows the left ventricle, left atrium, and left ventricular outflow tract. Useful for evaluating aortic valve and root.
- 4CH: Visualizes all four chambers: left atrium, left ventricle, right atrium, and right ventricle. Helps in assessing overall cardiac function.
- LVOT: Focuses on the area where the left ventricle ejects blood into the aorta. Critical for evaluating aortic stenosis or regurgitation.
- RVOT: Highlights the pulmonary valve and the path of blood from the right ventricle to the lungs. Important in conditions like pulmonary hypertension.
- SA: Series of cross-sectional images of the heart from apex to base, commonly used for volumetric analysis and observing physiological variations.

### 2.2 Model Details

This study compares a Caption transformer model and ClipCap model for the task of image labelling, to evaluate performance differences under a contained experimental setup. Although both the approaches use a transformer as the skeleton, it differs in the way they are trained. While the Caption transformer was trained from scratch, the same training data was used to fine-tune ClipCap. The hyperparameters, like the learning rate, batch-size were tuned to achieve the desired output, based on the results of the validation set.

#### End to end Transformer Network

This network uses a Vision Transformer (ViT) Network, which processes images the same way as a language transformer processes sentences. To obtain visual features, the input image is divided into many small patches and then passed to the encoder, containing multiple multi-head self-attention layers followed by positional feed forward layer. This acts as one of the inputs to the decoder along with the word embedding. The decoder consists of stacked masked multi-head self-attention sublayer followed by a multi-head cross attention sublayer and a positional feedforward sublayer [7]. Figure 2 gives the architectural overview of the end-to-end transformer network which is referred to as Caption Transformer.

**Figure 2:**
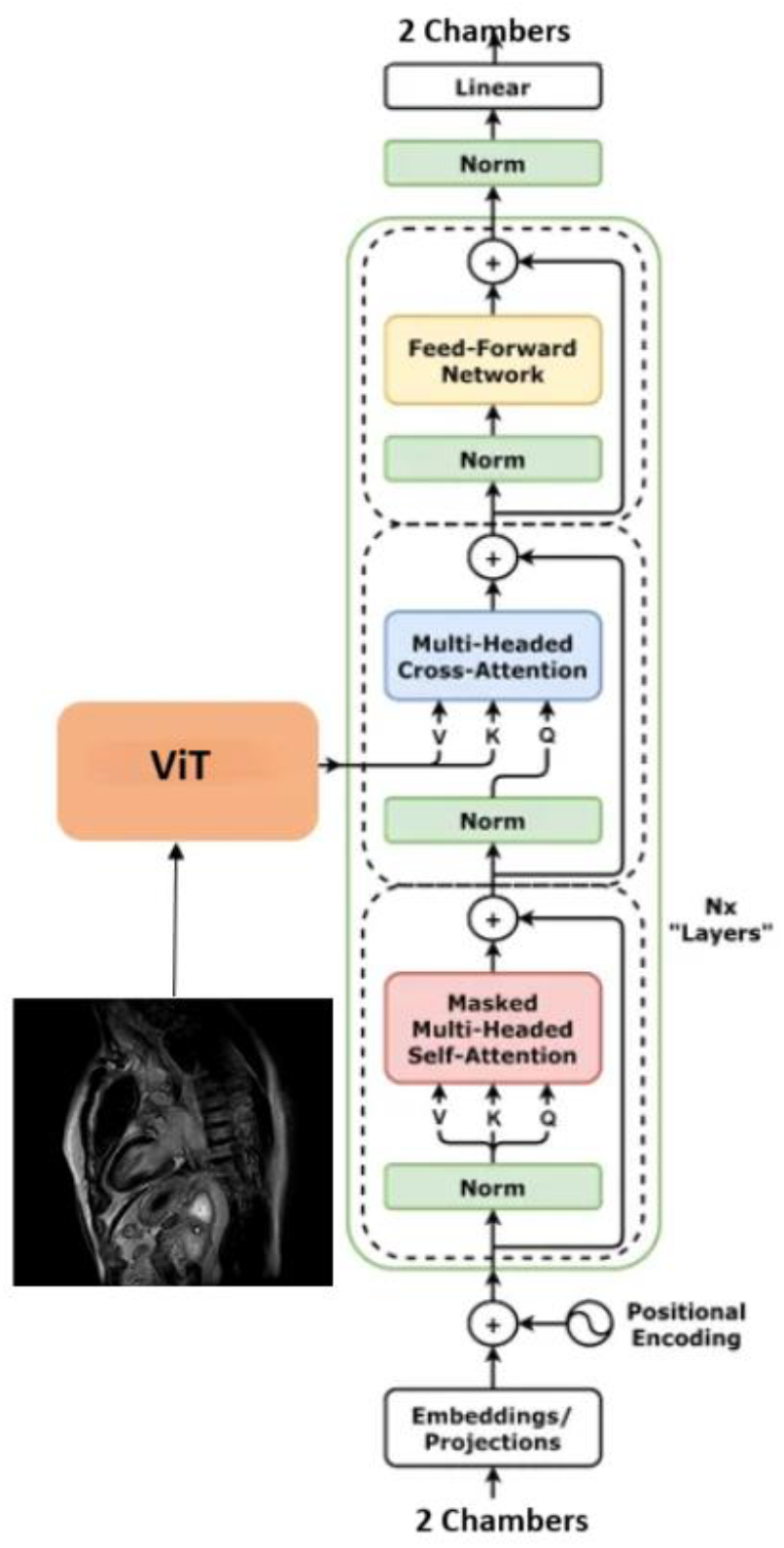
Architecture of end-to-end transformer

#### Contrastive Language-Image Pre-training for Captioning (CLIPCap)

CLIPCap has 3 main components, namely CLIP Encoder, Mapping Network and Language Model like GPT2 as in figure 3. The OpenAI’s CLIP model acts as an encoder, by generating image embeddings. It is a joint image and text embedding model and trained using 400 million image and text pairs in a self-supervised manner, to map the image and text into the same embedding space [8]. It uses a transformer network as a backbone and contains rich semantic features. The embeddings are then transformed into text-friendly embedding that serves as a prefix for the GPT model to generate the caption [9].

**Figure 3:**
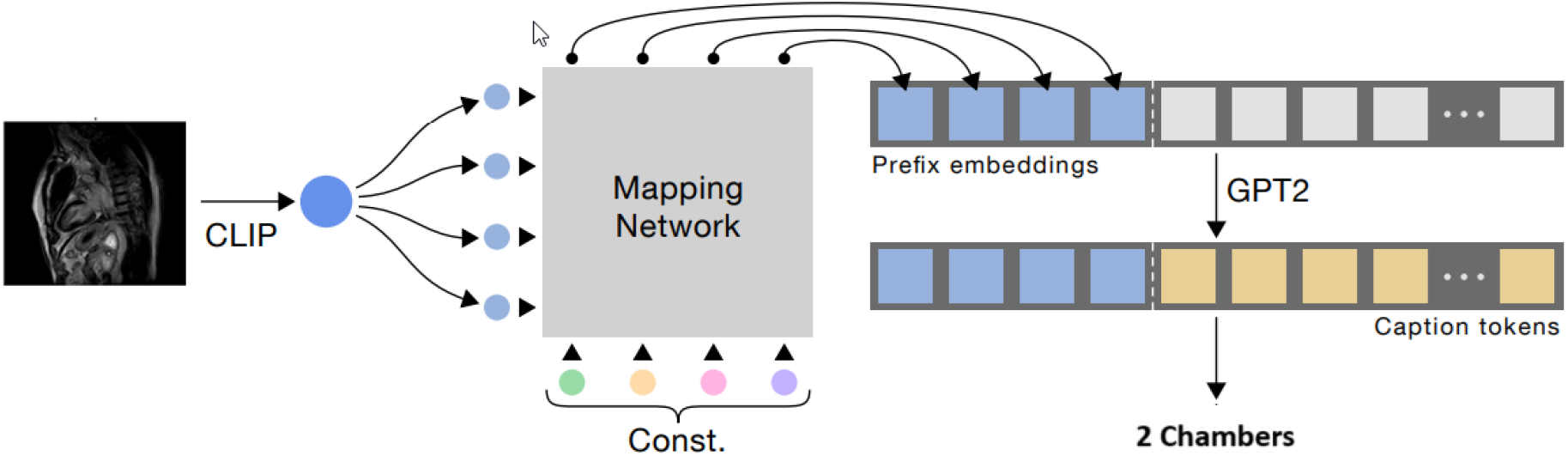
Architecture of CLIPCap model

**Figure 4:**
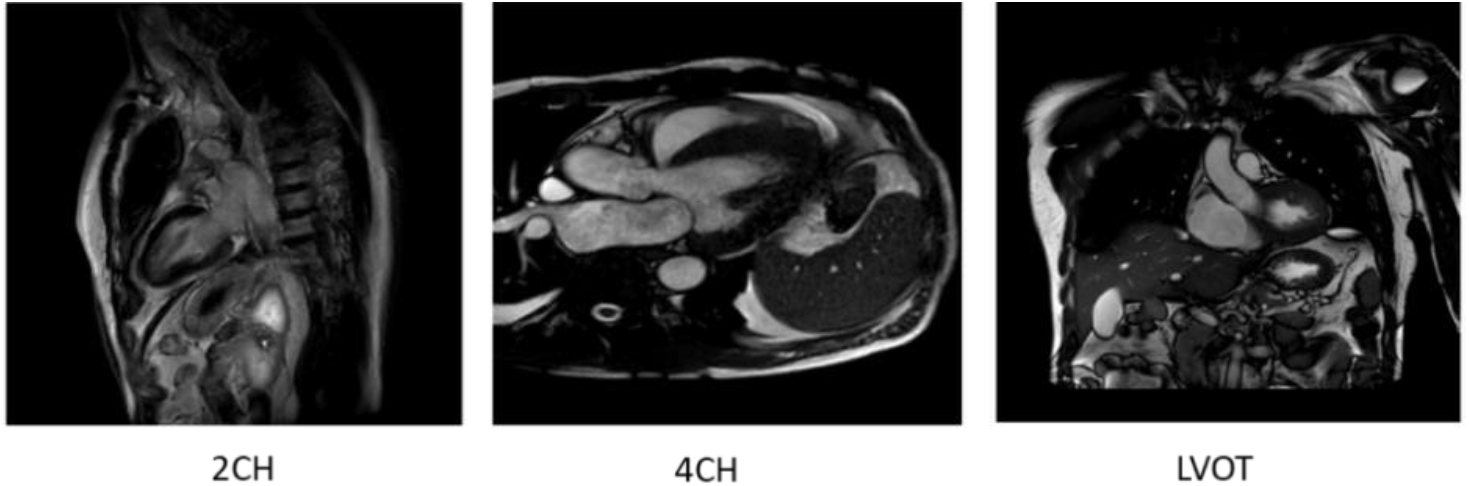
Example of correctly labelled class

Caption Transformer is a classic transformer-based image captioning model trained end-to-end on paired data whereas CLIPCap is more modular and data-efficient approach that maps CLIP image embeddings into GPT-2’s language space for caption generation.

The models were trained for 50 epochs using the Adam optimizer with a learning rate of 0.0001. To prioritize the most informative slices and reduce redundancy in cardiac magnetic resonance (MR) imaging, we applied a central-slice extraction strategy that retains only the middle one-third of slices from each volumetric acquisition. For a given stack comprising *N*slices, we computed the subset size *M* = ⌊*N*/3⌋ and selected a contiguous block of *M* slices centered within the stack (indices ⌊(*N* − *M*)/2⌋to ⌊(*N* − *M*)/2⌋ + *M* − 1). This approach emphasizes the anatomically relevant mid region while discarding basal and apical slices, which often exhibit partial coverage or increased variability. By focusing on the most representative portion of the cardiac volume, this method improves inter-subject comparability, reduces storage and computational burden.

## 3. Results

Comparative evaluation of ClipCap and Caption transformer model demonstrated a clinically meaningful performance advantage of ClipCap across both global and class-specific metrics. The image with the correct label in shown in figure4. ClipCap achieved an overall accuracy of 0.98, substantially exceeding the accuracy of 0.68 obtained by the transformer model in proving appropriate orientation labels to the MR Cardiac volumes. From a clinical perspective, this large performance gap indicates a significantly lower risk of mislabeling the orientation when using ClipCap, which is critical for downstream diagnostic interpretation and automated workflow integration.

At the class level, CLIPCap consistently delivered high precision and recall across all cardiac views, reflecting reliable identification with minimal false positives and false negatives as depicted in figure 5. In the 2CH view, CLIPCap achieved near-perfect precision (1.00) and high recall (0.95), resulting in an F1-score of 0.98. In contrast, CPTR exhibited reduced precision (0.80), indicating a higher false-positive rate that could lead to incorrect clinical interpretation despite comparable recall.

**Figure 5:**
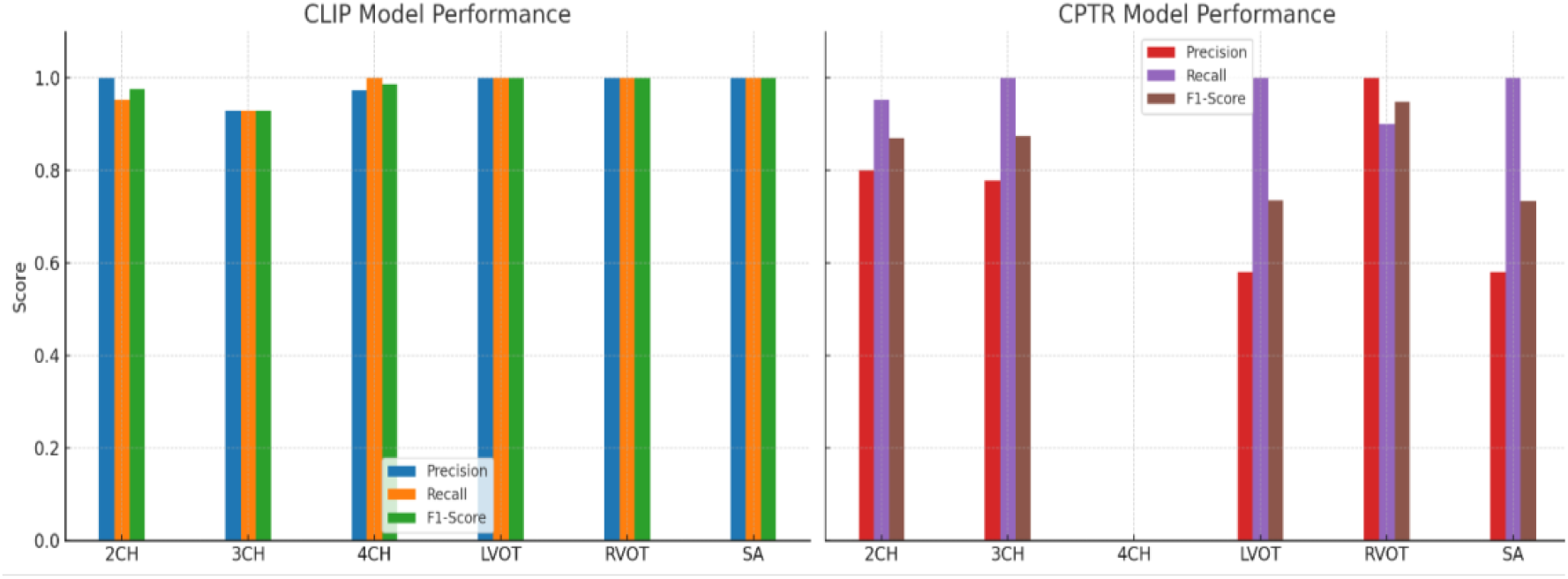
Comparison of Class-wise metrics: ClipCap Vs end-to-end transformer model

For the 3CH view, CLIPCap maintained a balanced and stable performance (precision, recall, and F1-score all 0.9286), whereas CPTR’s perfect recall (1.0000) was offset by lower precision (0.7778). This imbalance suggests that CPTR tends to overpredict this class, which may reduce clinical trust in automated outputs due to increased false alarms.

A particularly critical finding was observed in the 4CH class, which is of high clinical importance, as it provides long-axis plane that shows all 4 chambers of the heart simultaneously. CLIPCap demonstrated excellent performance with an F1-score of 0.9863, whereas CPTR failed to correctly identify any samples. Such a complete mislabeling represents a clinically unacceptable outcome, as mislabeling of the 4CH view could severely compromise automated cardiac assessment pipelines.

Similarly, for the LVOT view, CLIPCap achieved perfect classification across all metrics, supporting its reliability in identifying anatomically complex views. CPTR, however, exhibited moderate performance (precision = 0.5806, F1-score = 0.7347), primarily due to a high number of false positives, which could lead to inappropriate downstream measurements or diagnostic errors.

In the RVOT class, although both models performed well, CLIPCap again achieved perfect precision and recall, while CPTR showed a slight reduction in recall (0.9000). Even modest recall degradation in clinically relevant views can translate to missed cases in real-world screening scenarios.

Finally, for the SA class, CLIPCap maintained flawless performance, whereas CPTR’s lower precision (0.5789) and F1-score (0.7333) reflected substantial false-positive activity, undermining its clinical reliability.

Overall, CLIPCap demonstrated not only superior accuracy but also greater consistency and robustness across all cardiac views, minimizing clinically significant errors such as false negatives in essential views (e.g., 4CH) and excessive false positives in others (e.g., LVOT and SA). These characteristics are particularly important in clinical settings, where model stability and predictable performance are prerequisites for safe deployment. Consequently, CLIPCap represents a more clinically reliable and robust solution for automated cardiac view classification compared to the end-to-end transformer model.

## 4. Discussion & Conclusion

The present study provides a systematic comparison between a conventional caption transformer model and the CLIPCap architecture for automated cardiac MRI view labelling. Our findings demonstrate a clear and consistent performance advantage of CLIPCap across all cardiac orientations, both in terms of accuracy and class-specific metrics. The result supports our initial hypothesis that leveraging CLIP’s large scale and semantically rich image-language embeddings would confer improved generalization and robustness compared to a transformer model trained exclusively on a domain-specific dataset.

Across the six cardiac views, CLIPCap achieved near-perfect precision, recall and F1-scores in multiple classes (LVOT, RVOT, and SA), with an overall accuracy exceeding 98%. The caption transformer attained an overall accuracy of 68% with particularly poor performance in clinically relevant orientations such as 4CH view, which is crucial in assessing cardiac morphology, cardiac function and tissue characterization. Cardiac orientation labels serve as inputs in downstream automated analysis pipelines, such as chamber segmentation and functional assessment.

Misidentification of the views can directly impact these workflows, leading to inappropriate model selection and inconsistent clinical measurements. CLIPCap’s consistently high precision and recall reduce these risks and improve trustworthiness in a clinical deployment scenario, particularly in high-throughput environments where manual review of every scan is impractical.

This discrepancy suggests that caption transformer struggled to develop sufficient discriminative feature representation from the relatively limited training dataset. In contrast, CLIPCap benefited substantially from CLIP’s pre-trained image encoder, therefore bringing broader generalization capability into downstream tasks. The performance gap between the two architectures further reflects additional advantages of using CLIPCap. The richness of CLIP’s image-text embedding space is advantageous for medical images that needs eye to detail at a clinical level, which can be achieved by fine tuning the model with medical dataset. Second, the lightweight mapping network can converge efficiently on smaller datasets. These observations emphasize the fact that CLIPCap better suits scenarios where the data required for model development is limited.

The limitation worth noting is that the CLIPCap model being trained on millions of image-text pairs is architecture and parameter heavy. This may cause hindrance in the end deployment context, due to need for large infrastructure.

However, we expect that the CLIPCap model can be easily scalable/extended to other labeling such as different organ and different disease types with few shot learning. Proposed model can also be used in whole body CT or MR volumes to detect and label sub volume containing specific organs, example liver, lung, colon, etc. Identifying and labelling the sub volumes help in the selection of organ specific AI models for further processing. Example, after labelling the slices containing liver, the label can further be used to select Couinaud segmentation algorithm, (division of liver into 8 segments). In a similar way, once the model labels the slices with lung, this can be used to select the AI models that does lung lobe segmentation, lung nodule detection etc.

In conclusion, the study highlights the advantages of leveraging large-scale multimodal pretrained models for medical imaging tasks and demonstrates that CLIPCap provides a robust, efficient, and clinically reliable method for cardiac MRI view labelling. While further validation on larger and more diverse datasets is necessary, the findings underline the potential of contrastive vision– language models to enhance efficiency and consistency in clinical workflows.

## Data Availability

All data produced in the present study are available upon reasonable request to the authors and are subject to consent from institutional board

## 5. Acknowledgement

I would like to thank all co-authors for their careful review of the manuscript and for providing valuable comments and constructive feedback that helped strengthen the scientific content and presentation of this work. Their insights and suggestions significantly contributed to improving the quality and clarity of the manuscript.

## Notes

### Competing Interest Statement

The authors have declared no competing interest.

### Author Declarations

This study was conducted using a retrospective dataset of de-identified cardiac MR images. Data underwent a Privacy Impact Assessment and an Ethics Committee Clearance review, both of which were subsequently reviewed and approved by Philips Internal Committee for Biomedical Experiments

